# Cerebral Microvascular Injury in Severe COVID-19

**DOI:** 10.1101/2020.07.21.20159376

**Authors:** John Conklin, Matthew P. Frosch, Shibani Mukerji, Otto Rapalino, Mary D. Maher, Pamela W. Schaefer, Michael H. Lev, R. G. Gonzalez, Sudeshna Das, Samantha N. Champion, Colin Magdamo, Pritha Sen, G. Kyle Harrold, Haitham Alabsi, Erica Normandin, Bennett Shaw, Jacob E. Lemieux, Pardis C. Sabeti, John A. Branda, Emery N. Brown, M. Brandon Westover, Susie Y. Huang, Brian L. Edlow

## Abstract

**Importance:** Cerebral microvascular lesions are common in patients with severe COVID-19. Radiologic-pathologic correlation in one case suggests a combination of microvascular hemorrhagic and ischemic lesions that may reflect an underlying hypoxic mechanism of injury, which requires validation in larger studies.

**Objective:** To determine the incidence, distribution, and clinical and histopathologic correlates of microvascular lesions in patients with severe COVID-19.

**Design:** Observational, retrospective cohort study: March to May 2020.

**Setting:** Single academic medical center.

**Participants:** Consecutive patients (n=16) admitted to the intensive care unit with severe COVID-19, undergoing brain MRI for evaluation of coma or focal neurologic deficits.

**Exposures:** Not applicable.

**Main Outcome and Measures:** Hypointense microvascular lesions identified by a prototype ultrafast high-resolution susceptibility-weighted imaging (SWI) MRI sequence, counted by two neuroradiologists and categorized by neuroanatomic location. Clinical and laboratory data (most recent measurements before brain MRI). Brain autopsy and cerebrospinal fluid PCR for SARS-CoV-2 in one patient who died from severe COVID-19.

**Results:** Eleven of 16 patients (69%) had punctate and linear SWI lesions in the subcortical and deep white matter, and eight patients (50%) had >10 SWI lesions. In 4/16 patients (25%), lesions involved the corpus callosum. Brain autopsy in one patient revealed that SWI lesions corresponded to widespread microvascular injury, characterized by perivascular and parenchymal petechial hemorrhages and microscopic ischemic lesions.

**Conclusions and Relevance:** SWI lesions are common in patients with neurological manifestations of severe COVID-19 (coma and focal neurologic deficits). The distribution of lesions is similar to that seen in patients with hypoxic respiratory failure, sepsis, and disseminated intravascular coagulation. Collectively, these radiologic and histopathologic findings suggest that patients with severe COVID-19 are at risk for multifocal microvascular hemorrhagic and ischemic lesions in the subcortical and deep white matter.

**Key Points Section:** *Question:* What is the prevalence and pathophysiology of cerebral microvascular injury in patients with severe COVID-19?

*Findings:* In this retrospective cohort study of 16 patients undergoing MRI for neurologic complications of severe COVID-19, microvascular lesions were observed in 11 patients and showed an anatomic distribution similar to that seen in patients with hypoxic respiratory failure and sepsis. In one patient who died, brain autopsy revealed widespread microvascular injury, including perivascular microhemorrhages and microscopic ischemic lesions.

*Meaning:* Microvascular injury is common in patients with severe COVID-19. Radiologic-pathologic correlation, though limited to a single case, provides insights into possible mechanisms of injury.

## INTRODUCTION

Hundreds of thousands of people worldwide have survived severe coronavirus disease 2019 (COVID-19) since the beginning of the pandemic in December 2019.^1^ For survivors, recovery of brain function may lag behind that of other organs.^2^ Hypercoagulability, inflammation, and endothelial cell infection by severe acute respiratory syndrome coronavirus 2 (SARS-CoV-2) are potential mechanisms contributing to global brain dysfunction.^3^ Few brain imaging studies have been reported in patients with severe COVID-19,^4–10^ due to limited MRI access and difficulty in scanning critically ill patients. Fundamental questions about the pathogenesis of brain injury in patients with severe COVID-19 thus remain.

We investigated the imaging characteristics of COVID-19 brain injury by performing ultrafast high-resolution susceptibility-weighted imaging (SWI) MRI^11^ on a clinical MRI scanner located in our Neurosciences Intensive Care Unit (ICU). We focused on SWI because emerging evidence suggests that COVID-19 patients are at risk for microvascular lesions,^7–9,12^ and the SWI sequence provides optimal sensitivity for detecting microvascular lesions based upon their paramagnetic susceptibility effects in an MRI scanner’s magnetic field.^11^ In a cohort of 16 consecutively imaged critically ill patients with COVID-19, we tested the hypothesis that COVID-19 is associated with cerebral microvascular injury detectable by SWI. In one patient who underwent autopsy, we performed correlative radiologic-pathologic analyses and cerebrospinal fluid SARS-CoV-2 polymerase chain reaction (PCR) to explore the mechanisms of COVID-19 cerebral microvascular injury.

## METHODS

### Patients

Chart review identified all patients at our hospital who met the following inclusion criteria: (1) MRI performed on our Neurosciences ICU scanner between March 12 and May 14, 2020; (2) confirmed COVID-19 diagnosis by RT-PCR nasopharyngeal swab; and (3) severe COVID-19, as recently defined.^13^ Clinical characteristics and laboratory data were collected for all patients. This retrospective study was HIPAA compliant and was approved by the institutional review board with a waiver of consent.

### Imaging

All patients underwent brain imaging on a 3 Tesla Skyra MRI scanner (Siemens Healthcare) using an ultrafast 3D SWI sequence^11^ with the following parameters: TE/TR=21.5/40 ms, voxel dimensions=0.9×0.9×1.8 mm^3^, Wave-CAIPI acceleration *R*=6, total scan time 100 seconds. Conventional T1, T2, FLAIR, and diffusion-weighted images were also obtained. Two board-certified neuroradiologists (J.C., S.Y.H.) independently reviewed the SWI data. Foci of abnormal susceptibility signal were counted using an abbreviated version of the Microbleed Anatomical Rating Scale.^14^ The raters counted up to 10 lesions per anatomical location, after which they recorded “greater than 10”. We resolved discrepancies between the radiologist ratings by consensus.

### Brain Autopsy and Microscopy

An autopsy was performed for Patient 1, who died on hospital day 16. Routine microscopic analysis of the cerebral and cerebellar grey matter and white matter was performed after approximately two weeks of fixation in buffered formalin and standard paraffin embedding with Luxol & hematoxylin-and-eosin (LH&E) staining.

Immunohistochemical staining was performed with a Leica Bond III automated stainer, with mouse monoclonal antibodies directed against neurofilament (Dako M0762, 1:3200), CD3 (Leica PA0553, used as supplied), CD163 (Leica Cat #PA 0090, used as supplied) and CD68 (Biocare Cat # IP-033-AA, also used as supplied).

### SARS-CoV-2 Cerebrospinal Fluid PCR

For the Patient 1, extracted RNA was tested for the presence of SARS-CoV-2 using a probe-based RT-qPCR assay (see Supplementary Material for details).

### Statistical Analysis

Clinical and laboratory variables were compared between (1) patients with >10 SWI hypointense lesions, and (2) patients with <10 SWI hypointense lesions, using an unpaired t-test for normally distributed variables and Mann-Whitney U test for variables that were not normally distributed. P-values less than 0.05 were considered significant.

## RESULTS

### Susceptibility-weighted imaging

Sixteen patients with COVID-19 met inclusion criteria (Supplementary Table 1). All patients had acute respiratory distress syndrome (ARDS) and were mechanically ventilated at the time of MRI. MRI was performed because of unresponsiveness in 11 cases, and focal neurologic deficits in five cases. Punctate foci of abnormal susceptibility signal were identified in 11/16 cases (69%), and 8/16 (50%) had > 10 lesions (Figure 1, Supplementary Table 2).

**Figure 1:**
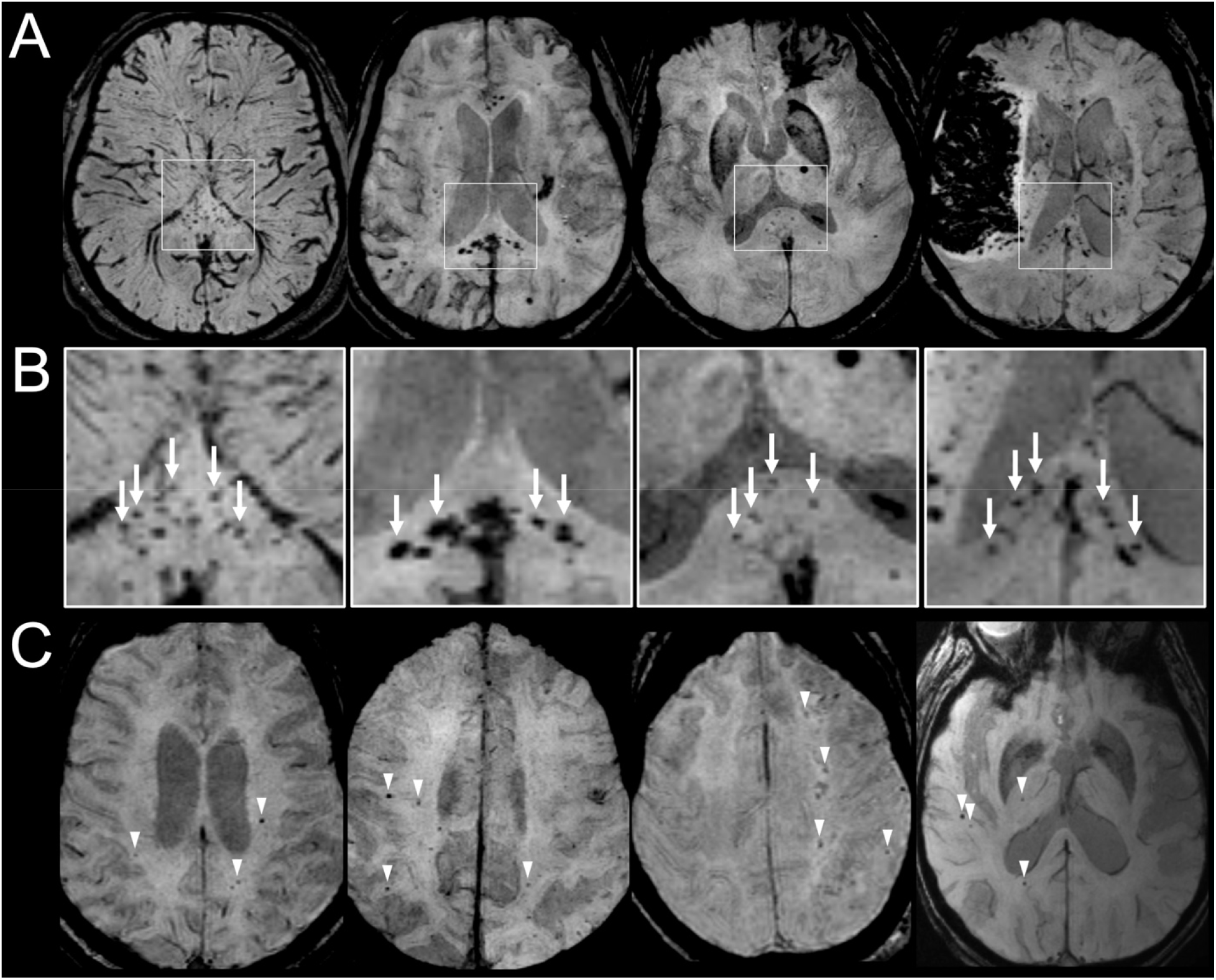
Representative susceptibility-weighted images from critically ill COVID-19 patients. (A) Four patients with susceptibility-weighted imaging (SWI) lesions including clustered lesions involving the corpus callosum. Patient 1 is shown in the first column. (B) Magnification views of the splenium of the corpus callosum for each patient shown in A, showing multiple SWI lesions (arrows). (C) Four patients with diffuse SWI lesions (arrowheads) involving the subcortical, periventricular and deep white matter, without involvement of the corpus callosum.

In 4/16 cases (25%), multiple clustered lesions involved the corpus callosum. In the remaining 4/16 cases with >10 lesions, the lesions showed a predilection for subcortical and deep white matter, with variable involvement of the brainstem and cerebellum. Additional MRI findings are summarized in Supplementary Table 2.

### Clinical and Radiologic-Pathologic Correlations for Patient 1

Patient 1 was a 57-year-old man with a history of asthma, obesity, and hyperlipidemia who tested positive for SARS CoV-2 by nasopharyngeal RT-PCR swab. His only COVID-19 related symptom on admission was shortness of breath, and he walked into the urgent care clinic. His hospital course was notable for hypoxic respiratory failure requiring mechanical ventilation and pupillary dilatation during weaning of sedation on hospital day 13. Head CT showed diffuse white matter hypoattenuation with preservation of gray-white matter differentiation and no downward herniation. Lumbar puncture was performed on day 14, and cerebrospinal fluid (CSF) analysis showed normal white blood cell count (1 cell/ul) and elevated total protein (181 mg/dL). RT-qPCR for the SARS-CoV-2 nucleocapsid protein targets (N1 and N2) performed on CSF was negative.

Brain MRI on day 15 revealed numerous SWI hypointense foci (Figure 1, left column), predominantly involving the subcortical and deep white matter, e.g., corpus callosum, internal capsules, and cerebellar white matter, as well as diffuse DWI and T2/FLAIR hyperintensity throughout the white matter. Life-sustaining therapy was withdrawn on hospital day 16, and he died shortly thereafter (see Supplementary Material for clinical details).

At autopsy, gross pathology revealed a diffusely edematous brain weighing 1410 grams (within normal range), with uncal and tonsillar herniation and diffuse discoloration of the gray-white matter junction. Numerous punctate hemorrhages were observed at the cortical gray-white matter junction and in the deep white matter.

Radiologic-pathologic correlations of the microvascular lesions are shown in Figure 2. Focused evaluation of the microvascular lesions showed microscopic foci of extravasated red blood cells associated with apparently necrotic blood vessels and foci of ischemic injury. The latter demonstrated significant loss, but not absence, of both axons and myelin, with occasional scattered axonal spheroids within the lesions and an associated macrophage/microglial response without other significant inflammatory cell components (Figure 3). Associated with the vessels nearest to the ischemic lesions, there was a moderate accumulation of microglia (CD163) and macrophages (CD68) (Figure 3). Detailed pathological findings are reported in the Supplementary Material.

**Figure 2:**
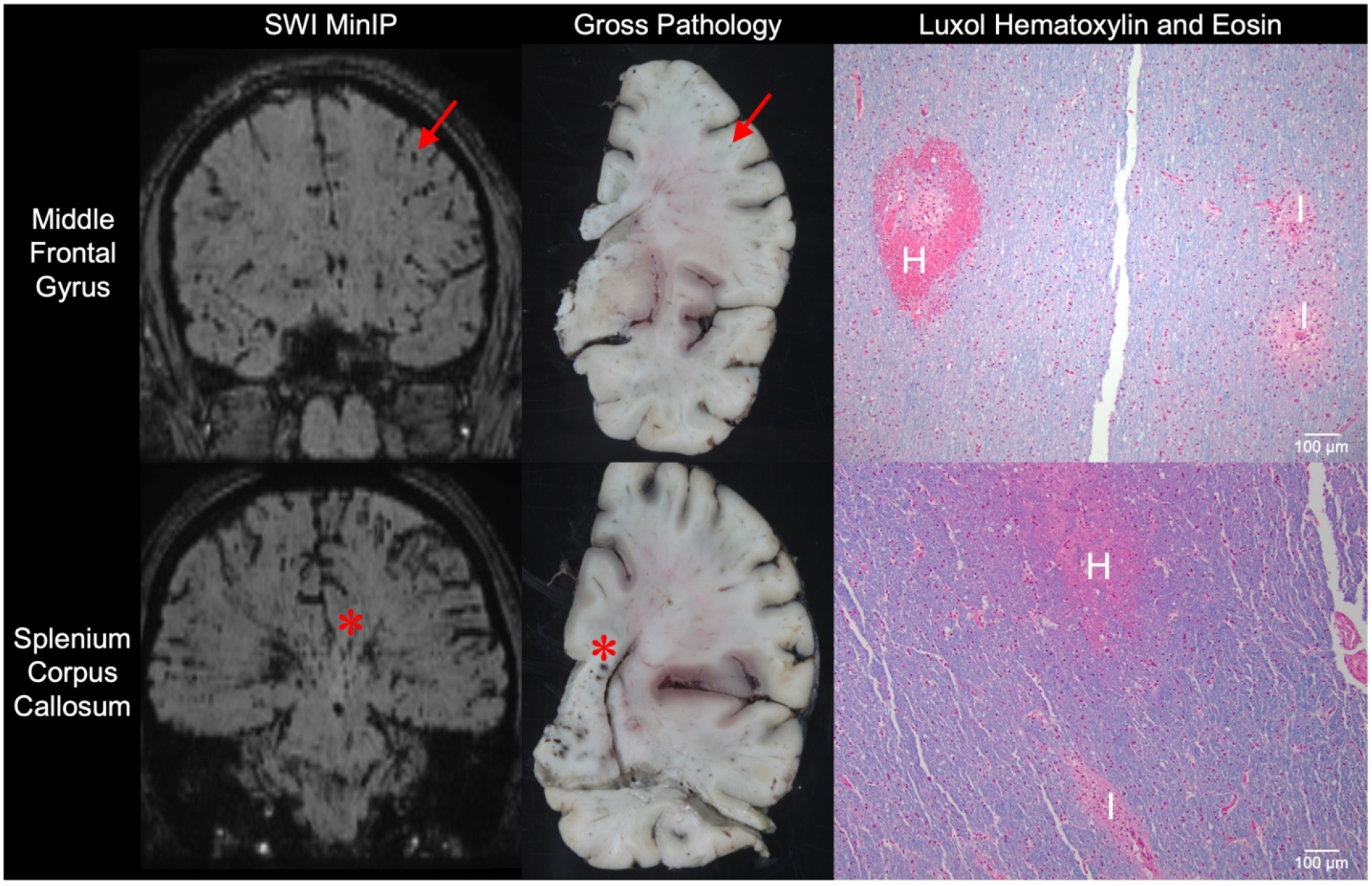
Radiology-pathology correlations in a patient who died from severe COVID-19. Susceptibility-weighted minimum-intensity projection images (SWI MinIP) are shown for the Patient 1 in the coronal plane at the level of the middle frontal gyrus (top row) and splenium of the corpus callosum (bottom row). Representative punctate hypointense foci are indicated by a red arrow (middle frontal gyrus) and asterisk (splenium of the corpus callosum). Gross pathologic analysis reveals corresponding lesions in coronal tissue slabs, as indicated by the red arrow and asterisk, respectively. On microscopic analysis of tissue stained with Luxol hematoxylin and eosin (LH&E), the lesions seen on SWI MinIP and gross pathology are characterized by microhemorrhages (H) and microscopic ischemic lesions(I).

**Figure 3.**
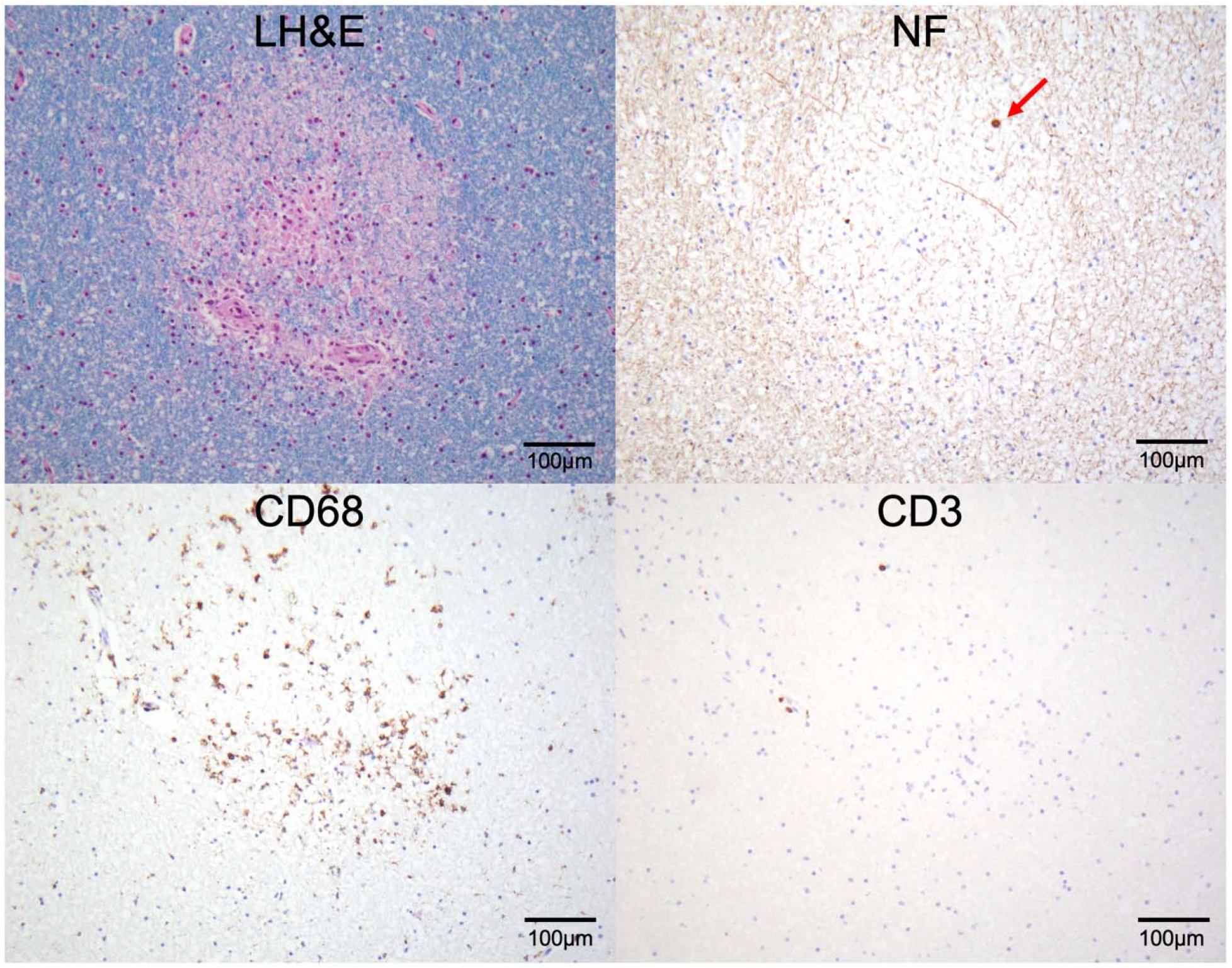
Pathology of ischemic lesions in a patient who died from severe COVID-19. Microscopic analyses of cerebellum at 200x magnification are shown for Patient 1. In the top left panel, a representative focus of microscopic ischemic lesion with significant loss, but not absence, of myelin is seen with Luxol hematoxylin and eosin (LH&E) stain. In the top right panel, neurofilament (NF) immunohistochemistry stain of the same lesion shows a significant loss, but not absence, of axons with rare axonal spheroids (red arrow). In the bottom left panel, a CD68 immunohistochemistry stain demonstrates aggregates of macrophages associated with the ischemic lesion. In the bottom right panel, a CD3 immunohistochemistry stain shows only rare CD3-positive T cells, indicating an absence of significant inflammation associated with the lesion.

## DISCUSSION

In this study of 16 critically ill patients with COVID-19 who underwent brain MRI because of persistent unresponsiveness (n=11) or focal neurologic deficits (n=5), we detected diffuse microvascular injury involving the subcortical and deep white matter in 69% of patients. Microvascular lesions manifested as punctate and linear hypointense foci on SWI, with a neuroanatomic predilection for the corpus callosum and the subcortical and deep white matter. There were no significant differences in clinical or laboratory variables between patients with and without SWI lesions. In one patient who died and underwent brain autopsy, the lesions showed mixed pathology: microhemorrhages and microscopic ischemic lesions. While the hemorrhagic lesions could be identified by SWI, no definitive imaging correlate to the microscopic ischemic lesions was observed, likely due to their small size relative to the spatial resolution of clinical DWI. Collectively, these findings suggest that cerebral microvascular lesions are common in severe COVID-19 patients who have neurologic deficits and that the pathogenesis of the microvascular lesions involves both hemorrhagic and ischemic etiologies.

Our correlative radiologic-pathologic findings add to the growing literature on the neurological effects of COVID-19 and shed new light on mechanisms of brain injury in severe COVID-19. The neuroanatomic distribution of the microvascular lesions, particularly the callosal and capsular predominance, has been reported as a rare complication of ARDS, high altitude exposure, and extracorporeal membrane oxygenation (ECMO) – all of which are associated with cerebral hypoxia.^15,16^ Endothelial cell infection and endotheliitis have been described in extracerebral organs of patients with COVID-19,^3^ presumably mediated via the ACE2 receptor, which is also found in brain endothelial cells.^17^ Our findings thus suggest a potential role for hypoxic microvascular injury or endothelial dysfunction in the pathogenesis of cerebral microvascular injury in severe COVID-19.

These observations, if replicated, have clinical relevance at both the individual patient and population levels. For individuals, detection of diffuse microvascular injury may be immediately actionable. Differentiating microhemorrhage from microthrombosis remains a diagnostic challenge because both appear hypointense on SWI.

Nevertheless, identification of SWI lesions could alter the risk-benefit analysis of anticoagulation, a particularly important consideration given the hypercoagulability observed in COVID-19 patients.^18^

At the population level, these imaging findings raise fundamental questions about neuroprotective strategies in COVID-19 patients undergoing mechanical ventilation for hypoxic respiratory failure. Could tracking serum-based biomarkers of bleeding, hypercoagulability, and COVID-related hyperinflammatory states or cytokine release syndrome^19^ identify patients at risk for cerebral microhemorrhage and ischemia? Could COVID-19-related hypoxia contribute to the pathogenesis of microvascular injury, as suggested by reports of patients with ARDS and “critical illness-associated cerebral microbleeds”?^15,16^ Finally, does cerebral microvascular injury contribute to the prolonged unresponsiveness being observed in survivors of severe COVID-19?^2,10^ All of these questions warrant urgent, systematic investigation to optimize neurologic outcomes in severe COVID-19.

## Data Availability

Available upon request.

## Acknowledgments

We thank the nurses, respiratory therapists, and MRI technologists who assisted in acquisition of the imaging data, and we thank the pathology technicians who assisted in the brain autopsy.

